# Germline Mutations Associated with Triple Negative Breast Cancer in US Hispanic and Guatemalan Women using Hospital and Community-Based Recruitment Strategies

**DOI:** 10.1101/2023.07.01.23292051

**Authors:** Jesica Godinez Paredes, Isabel Rodriguez, Megan Ren, Anali Orozco, Jeremy Ortiz, Anaseidy Albanez, Catherine Jones, Zeina Nahleh, Lilian Barreda, Lisa Garland, Edmundo Torres Gonzalez, Dongjing Wu, Wen Luo, Jia Liu, Victor Argueta, Roberto Orozco, Eduardo Gharzouzi, Michael Dean

## Abstract

**Purpose:** Identify optimum strategies to recruit Latin American and Hispanic women into genetic studies of breast cancer. We evaluated hospital and community-based recruitment strategies.

**Methods:** We used targeted gene sequencing to identify mutations in DNA from unselected Hispanic breast cancer cases from community and hospital-based recruitment in the US and Guatemala.

**Results:** We recruited 287 Hispanic US women, 38 (13%) from community-based and 249 (87%) from hospital-based strategies. In addition, we ascertained 801 Guatemalan women using hospital-based recruitment. In our experience, a hospital-based approach was more efficient than community-based recruitment. In this study, we sequenced 103 US and 137 Guatemalan women and found 11 and 10 pathogenic variants, respectively. The most frequently mutated genes were *BRCA1, BRCA2, CHEK2*, and *ATM*. In addition, an analysis of 287 US Hispanic patients with pathology reports showed a significantly higher percentage of triple-negative disease in patients with pathogenic mutations (41% vs. 15%). Finally, an analysis of mammography usage in 801 Guatemalan patients found reduced screening in women with a lower socioeconomic status (P<0.001).

**Conclusions:** Guatemalan and US Hispanic women have rates of hereditary breast cancer mutations similar to other populations and are more likely to have early age at diagnosis, a family history, and a more aggressive disease. Patient recruitment was higher using hospital-based versus community enrollment. This data supports genetic testing in breast cancer patients to reduce breast cancer mortality in Hispanic women.

## Introduction

Breast cancer is the most prevalent cancer among women worldwide (1), with a mortality rate of 30% in 2020. Breast cancer incidence is higher in countries with higher socioeconomic status, but mortality rates are lower. In contrast, in low-and-middle-income countries (LMIC), breast cancer has a lower prevalence rate but a higher mortality rate among women (2017 GLOBOCAN). This disparity is likely due to treatment availability and early detection in higher-income countries. In 2020 breast cancer accounted for 24% of cancer cases among women in Guatemala [1]. Whereas in women in the United States (US), in 2019, 30% of newly diagnosed cancer cases were breast cancer [2].

Most genetic and epidemiological studies in the US have focused on populations of European descent [3]. Although there are programs centered on Hispanic populations, there are obstacles to the recruitment of minority women in research, such as community engagement, language, and cultural barriers [4]. Latin American and US Hispanic women are underrepresented in genetic breast cancer studies. Failure to detect *BRCA1* and *BRCA2* mutations can delay diagnosis and treatment. Increased availability and affordability of next-generation sequencing and annotation of genetic variants would aid in identifying inherited mutations among Hispanic populations and reduce mortality. This study aims to analyze high penetrance breast cancer genes with pathogenic mutations in Hispanic women in US and Guatemalan women. Furthermore, we highlight the importance of patient recruitment strategies to increase the Hispanic women’s participation in genetic studies.

## Methods

### Patient recruitment in Guatemala

In Guatemala City, we recruited patients from the Hospital General San Juan de Dios (HGSJDD) and the Instituto Nacional de Cancerologia (INCAN). Both Hospitals serve the entire Guatemalan population, with patients referred by primary care physicians. Most breast cancer patients treated at INCAN were older than 40 (84%) and lived in the capital city, indicating a higher SES. Furthermore, younger patients came from regions west of Guatemala City (22%).

The questionnaire for Guatemalan patients included age at the time of diagnosis, demographics, reproductive history, cooking on a wood stove (a measure of SES), and family history of breast cancer.

### Patient recruitment in the US

Community recruitment across the US was conducted from July 2011 to August 2016. The study was published online in ClinicalTrials.gov and advertised on social media. we also recruited patients at community centers, such as Nueva Vida, Baltimore and Richmond (https://www.nueva-vida.org/), the Avon Breast Cancer Walk, Washington, DC (https://www.avon.com/breast-cancer-crusade), and the Oklahoma Latino Community Development Agency. In addition, we sent letters to oncologists in Southern Florida.

In Texas, we recruited subjects from the Texas Tech University Health Sciences Centers at Lubbock and El Paso. We invited women receiving care for current or previous breast cancer diagnoses to participate in the study. All US and Guatemala patients were administered a questionnaire in Spanish or English. The questionnaire for participants included age of diagnosis, demographics, reproductive history, socioeconomic status, and relevant family history of breast cancer. The questionnaire was identical for Guatemalan and US subjects. (**Appendix A**).

### IRB Approval and patient consent

In Guatemala this study was conducted at the Hospital General San Juan de Dios (HGSJDD) and the Instituto de Cancerología (INCAN) in Guatemala City. The Research Ethical Committees of each institution approved the protocol, and the study was determined exempt from institutional review board (IRB) approval by the NIH Office of Human Studies Research. Women attending either of these hospitals for their breast cancer diagnostic biopsies were invited to participate and gave written informed consent. Two 5 ml tubes of blood were collected and frozen at -20°C as well as a tumor biopsy stored in 0.5 ml of RNAlater solution at -20°C. Trained interviewers administered an approved questionnaire including reproductive history, family history of cancer, and socioeconomic data.

In the U.S IRB protocol was approved by the NCI IRB and is listed in clinical trials.gov (https://www.clinicaltrials.gov/ct2/show/NCT01251900) (NCT01251900)

### Sequencing

We used targeted sequencing to identify mutations in blood DNA from unselected Hispanic breast cancer cases from community recruitment and from two hospitals each in Texas and Guatemala. A total of 137 blood samples from INCAN, HGSJDD, and 96 blood samples from Texas (El Paso and Lubbock) were sequenced on the NOVASeq from Illumina with the Paired-end 200bp strategy [5]. Briefly, blood DNA (200 ng) were used to produce an adapter-ligated library the Kapa HyperPlus Kit (Roche, Indianapolis, IN) using xGen Dual Index UMI Adapters (IDT, Coralville, IA) according to Kapa-provided protocol. The resulting post-capture enriched multiplexed sequencing libraries were loaded on a NovaSeq 6000 (Illumina, San Diego, CA) and paired-end sequencing was performed using read lengths of 2×150bp to an average coverage of 50x.

### Mutation classification

We analyzed high protein truncating variants in the genes *BRCA1, BRCA2, PALB2, CHEK2*, and *ATM*, as well as pathogenic missense variants in *TP53, BARD1, RAD51C*, and *RAD51D* [6]. All genes are listed in the **Supplemental Table. 1**

We annotated variants using SNPNexus for targeted breast cancer genes. We performed manual validation using Integrative Genomics Viewer (IGV) (https://igv.org/app). Then placed the variants into three categories pathogenic, VUS, and benign. Pathogenic mutations were further confirmed using ClinVar (https://www.ncbi.nlm.nih.gov/clinvar/) and Varsome (https://varsome.com/).

### Breast Cancer subtypes in US Hispanic Women

Hormone receptor data was only available in the US Hispanic subjects. We characterized the four breast cancer subtypes as follows: 1) luminal A as estrogen receptor (ER) positive, progesterone receptor (PR) positive, HER2 receptor-negative, and low Ki 67. 2) Luminal B subtype ER positive, PR positive, HER2 +/-, and high Ki67 count. 3) HER2+ subtype is ER negative, PR negative, and HER2 positive regardless of Ki67 count. 4) Triple-negative breast cancer is ER, PR, and HER2 receptor-negative. We combined ER and PR positive, HER2 negative tumors without Ki67 data into a luminal A/B group [7].

### Mammography Screening

Self-reported mammography screening was available for both Guatemala and US Hispanic women. We calculated active mammography screening for patients over the age of 40. We determined the difference between the age of diagnosis and age at the first mammogram. If the difference was two years or greater, we classified the subject as receiving active screening.

Patients with a first mammogram less than two years before diagnosis were classified as unscreened.

### Socioeconomic Status (SES)

We used self-reported income brackets in the US to estimate socioeconomic status (SES). In Guatemala, SES is difficult to determine directly due to the lack of job security. As a proxy, we used cookstove type as an indicator of SES due to the known association between wood-burning stoves and poverty in Guatemala, particularly among indigenous Mayans (https://doi.org/10.1186/ISRCTN29007942, guatemalastoveproject.org/). We confirmed that wood cookstove use is associated with Indigenous American ancestry (**Supplemental Figure 1**).

### Statistics

We used a two-proportion Z-test (two-tailed) to assess the difference in the percentage of patients with a family history of breast cancer, contraception use, and parity. Second, we used an unpaired t-test with Welch’s correction to ascertain the relationship between breast cancer subtypes in US patients with pathogenic and non-pathogenic mutations. Third, a Chi-squared test with Yates correction examined the relationship between socioeconomic status (cookstove type) and mammogram screen usage. Finally, Fisher’s exact test assessed the relationship between Guatemalan women’s cookstove use and Indigenous American ancestry. In all calculations, a p-value of 0.05 or less was deemed significant.

## Results

### Patient Recruitment

To analyze the characteristics of breast cancer in Hispanic patients in the US and Guatemala we recruited women through community-and hospital-based strategies. Our current study ascertained 103 US Hispanic women from hospital-based recruitment in TTUHSC El Paso and Lubbock. We ascertained 137 women from the INCAN and HGSJDD hospitals in the Guatemala cohort. In **Figure 1**, we indicate the number of women sequenced for germline mutations and report the total of each with and without pathogenic mutations. In a previous study in the US, we described *BRCA1* and *BRCA2* mutations in 184 women [8]. Overall, in the US, we recruited 38 (13%) women through community-based recruitment and 249 (87%) from two hospitals in Texas (**Table 1**). In addition, we recruited a total of 801 patients using hospital-based recruitment in Guatemala City; 137 from our current study and 664 from a previous study [5].

**Table 1.** Demographics of US and Guatemalan women.

|  | Total Guatemala n=801 |  | Total US n= 287 |  | *p*-value |
| --- | --- | --- | --- | --- | --- |
|  | Median (IQR) | Range | Median (IQR) | Range |  |
| Age at Diagnosis | 49 (41-61) | 19-93 | 52 (43-60) | 23-81 | 0.57 |
| Age at first mammogram | 47 (40-59) | 15-91 | 40 (35-45) | 17-75 | <0.0001 |
| Age at menarche | 13 (12-14) | 9-24 | 13 (11-14) | 8-18 | <0.0001 |
| Age at 1st pregnancies | 21 (18-25) | 14-43 | 21 (18-24) | 13-42 | 0.37 |
| Number of pregnancies | 3.0 (2.0-5.0) | 0-19 | 3.0 (2.0-4.0) | 0-10 | <0.0001 |
| Number of children | 3.0 (2.0-4.0) | 0-19 | 3.0 (2.0-4.0) | 0-8 | 0.0004 |
| Breast feeding | 90% |  | 50% |  | <0.0001 |
| Family history | 14% |  | 40% |  | <0.0001 |

Analysis of patient demographics and risk factors known to be associated with the development of breast cancer. We report median, interquartile ranges (IQR), and *p*-values for all Guatemala and US women. The family history reported is solely for family members with breast cancer.

**Figure 1.**
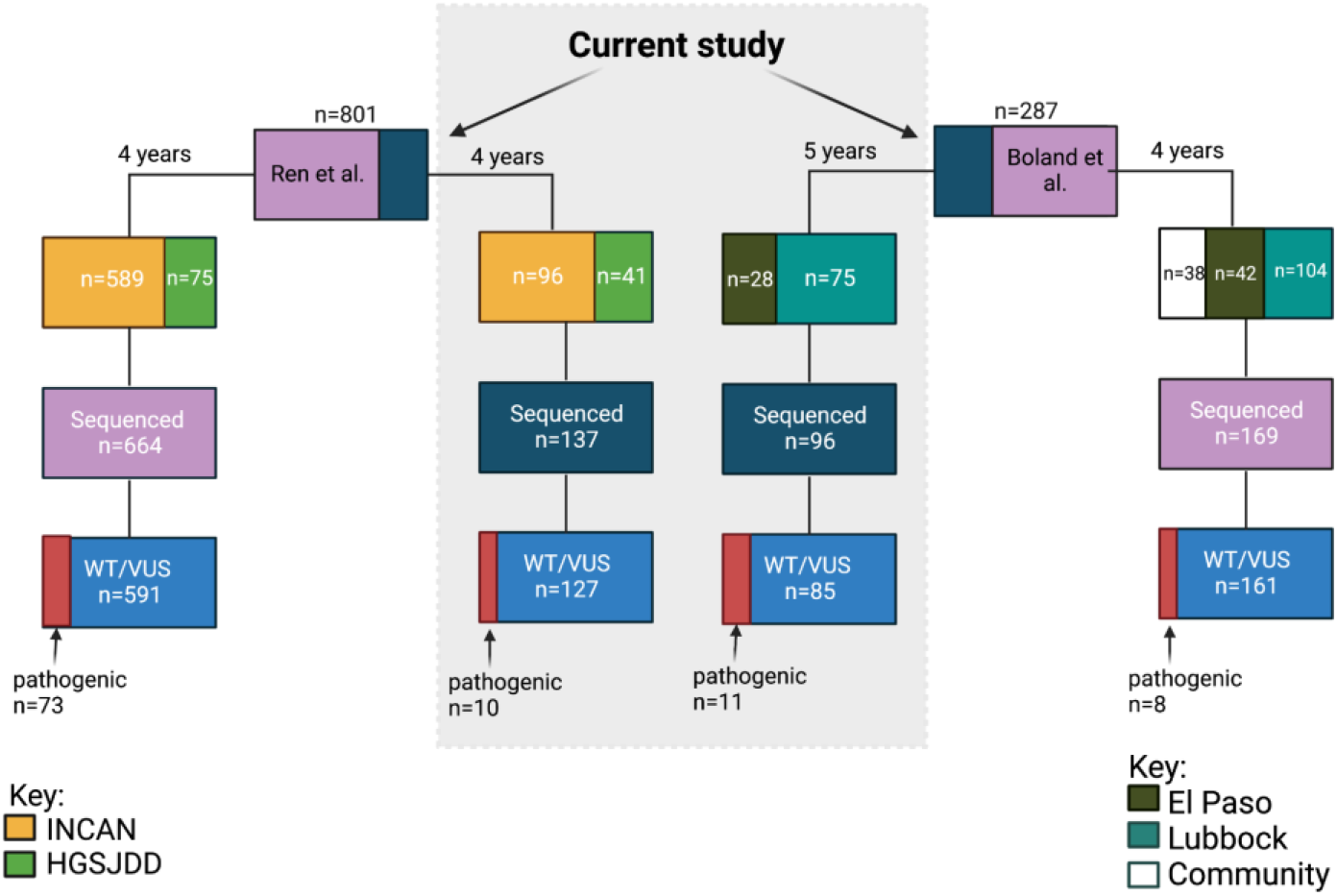
Recruitment of US Hispanic and Guatemalan women. We show the patient distribution of subjects in Guatemala and the US. There was a total enrollment of 801 unselected Guatemalan women through hospital-based recruitment over 5.25 years. Enrollment in the US occurred over 9 years with 287 unselected women, 38 from community recruitment, and 249 from a hospital-based approach. We report the US and Guatemalan patients in this study within the gray box. Partial data on additional subjects from the two cohorts have been previously published [5, 8].

### Demographics

In breast cancer, age is one of the most significant risk factors, as the development of the disease increases after the fourth decade of life [2]. The median age at diagnosis for Guatemalan women was 49 years old, and for US Hispanic women 52 years old, with no significant difference between these two groups. Early age at menarche increases a woman’s risk of developing breast cancer due to earlier exposure to estrogen [9]. Guatemalan and US Hispanic women had the same median age of menarche, 13 years old. However, the mean age in Guatemala was 13.2, and in the US was 12.6 (p<0.0001) (**Table 1**). Decreased estrogen exposure can reduce a woman’s risk of developing breast cancer, and reproductive factors that decrease risk include higher parity, lower age at first birth, and breastfeeding [9]. Both groups’ median age at first pregnancy was 21, with no differences between Guatemalan and US women. The mean of live births in Guatemala, 3.3 was significantly higher than in US women, 2.8 (p=0.0004). Early diagnosis of breast cancer can ultimately result in an improved prognosis and reduced incidence of metastatic cancer. Currently, the US guidelines recommend mammography screening for women at 50 years old, and women with a family history of breast cancer are recommended mammography screening at 40 years old [10]. In our analysis, the median age of Guatemalan women at their first mammogram was 47 years old, significantly higher (p<0.0001) than US Hispanic women with a median age of 40. Therefore, our data suggest that US Hispanic women, in our study, received mammography screening consistent with current guidelines.

### Analysis of pathogenic mutations

We previously published pathogenic mutations in 73 out of 664 Guatemalan patients and 10 out of 96 patients from the US Latina population. In this study we sequenced an additional 137 Guatemalan breast cancer cases and identified 10 pathogenic mutations in high penetrance genes (**Table 2)**. We identified three additional cases of the c.212+G>A founder mutation in *BRCA1*. Furthermore, we present current data on 96 additional US Hispanic patients, identifying 11 more pathogenic mutations in high and moderate penetrance genes (**Table 2)**. We identified a carrier of the c.68_69delAG Ashkenazi Jewish founder mutation in one patient from Texas, who had an early age diagnosis (40-45 years) and a family history of breast cancer. In the US combined data set, cases with pathogenic mutations had a significantly earlier age at diagnosis (41 vs. 52 years, P<0.0001) and were more likely to have a diagnosis before menopause. In addition, a higher percentage of mutation carriers had a relative with breast cancer (39% vs. 67%, P<0.01).

**Table 2.**
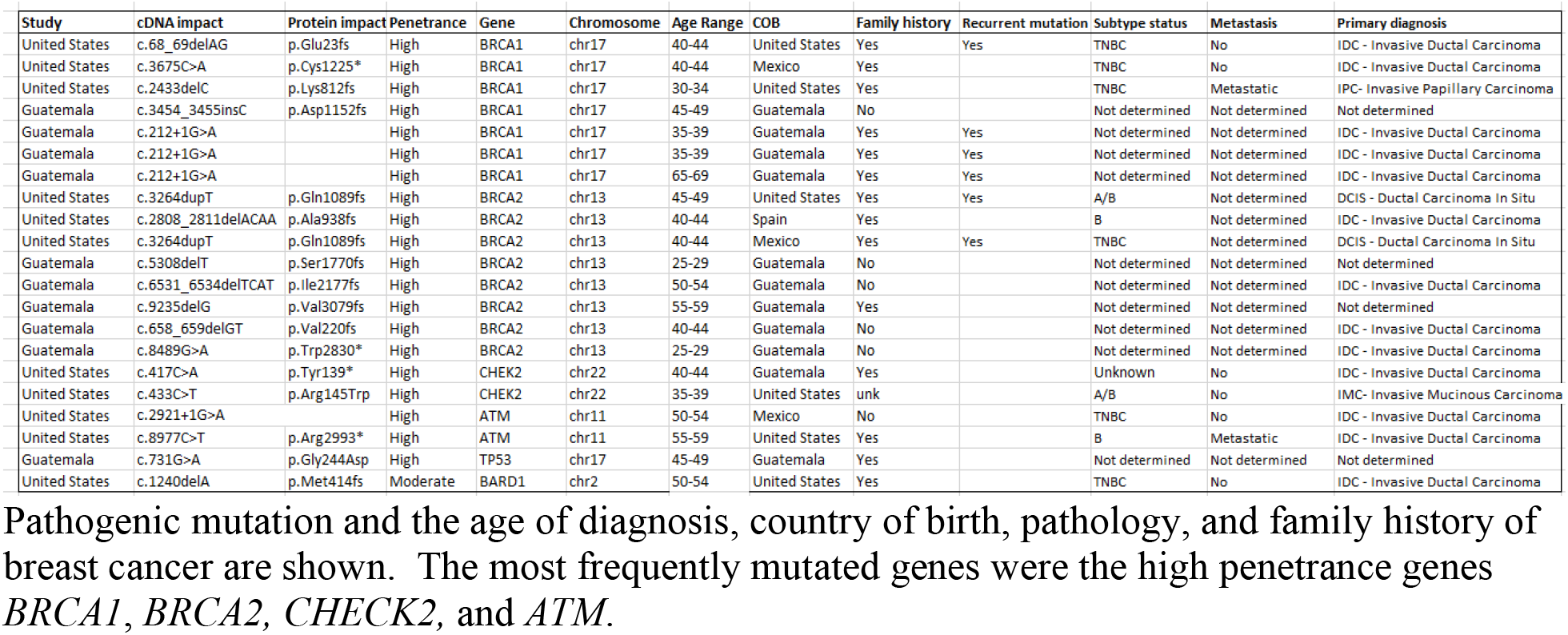
Pathogenic mutations include cDNA position, protein alteration, gene, and pathological status.

### Breast cancer subtypes

To determine if cases with a pathogenic mutation have a higher rate of triple-negative disease, we analyzed breast cancer subtypes in the entire US Hispanic population data set.

Patients with pathogenic mutations had a higher percentage of triple-negative disease (41% vs. 15%, P=0.9), and there were no HER2+ cases in this group (**Figure 2**). Therefore, US Hispanic women with pathogenic mutations have a higher frequency of aggressive breast cancer subtypes (TNBC) with poorer prognosis.

**Figure 2.**
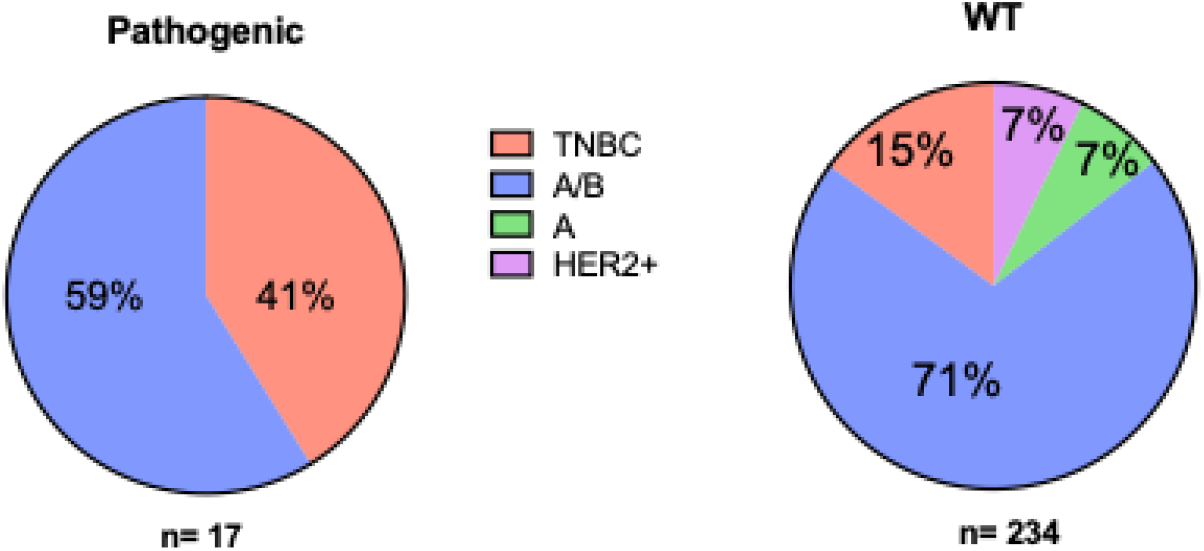
Breast cancer subtypes by mutation status. Analysis of breast cancer subtypes in 251 US patients with (n=17) and without pathogenic mutations (n=234). Subtypes were luminal A (green), luminal A/B (luminal A or B) (blue), HER2+ subtype (pink), and triple-negative (TNBC) (blue).

### Mammography screening

The American Cancer Society measures breast cancer screening rate by the percentage of women 40 and older who had a mammogram in the past two years (American Cancer Society). We sought to identify a connection between mammogram use and socioeconomic status. Self-reported mammography data was available for 611 Guatemalan women over 40. In total, 362 women (59%) indicated that they had received a mammogram, but only 249 (41%) had regular screening in the two years preceding their diagnosis (**Figure 3a**). Women in Guatemala did not report income; therefore, we used cooking with wood as a proxy for lower SES and for Indigenous American ancestry (**Supplemental Figure 1**). We found that mammography usage was less frequent in women cooking with wood, indicating that this group received inadequate breast cancer screening (P<0.0045).

**Figure 3.**
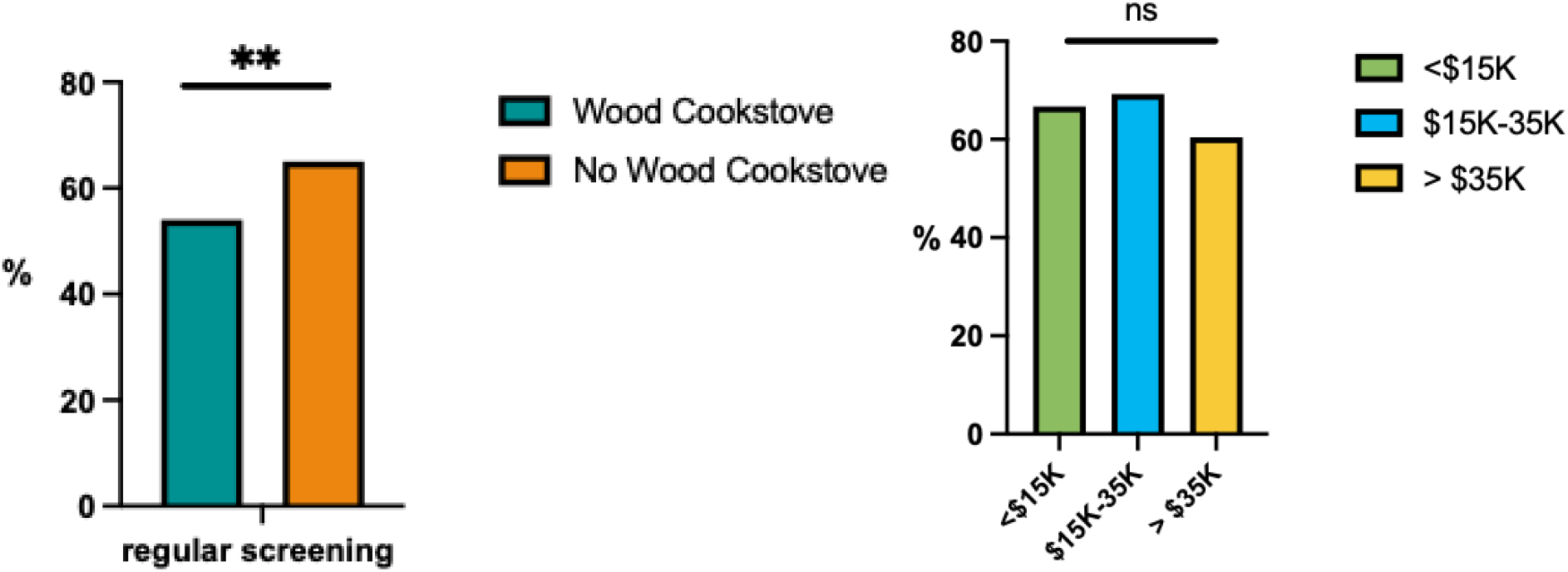
Mammography screening by socioeconomic status. **a**) Regular mammogram screening compared in women who do or do not use a wood cooking stove in Guatemala as a measure of SES. **b**) US women’s SES and screening regularity by income.

For women in the US, self-reported household income was available. We aggregated household income into three groups: less than $15,000, $15,000 to $35,000, and greater than $35,000. Mammography usage was not significantly different between SES groups in the US (**Figure 3b**), indicating that breast cancer screening is prevalent in US Texas women. The average household income in the US in 2021 was $70,784, whereas most women in our cohort were under the household median income. Therefore, despite the low socioeconomic status of the women in the US we studied, they are receiving regular mammography screenings.

### Metastasis in US women and SES

We compared income groups to determine if the prevalence of metastatic breast cancer is related to SES in US women. Between the three income subgroups, there was no significant difference in the percentage of metastasis (**Figure 4**). Therefore, women with an income less than $15,000 are being provided equal mammography screening as the other groups and are not developing a higher rate of metastatic disease, indicating there is no detectable health disparity for metastatic disease.

**Figure 4.**
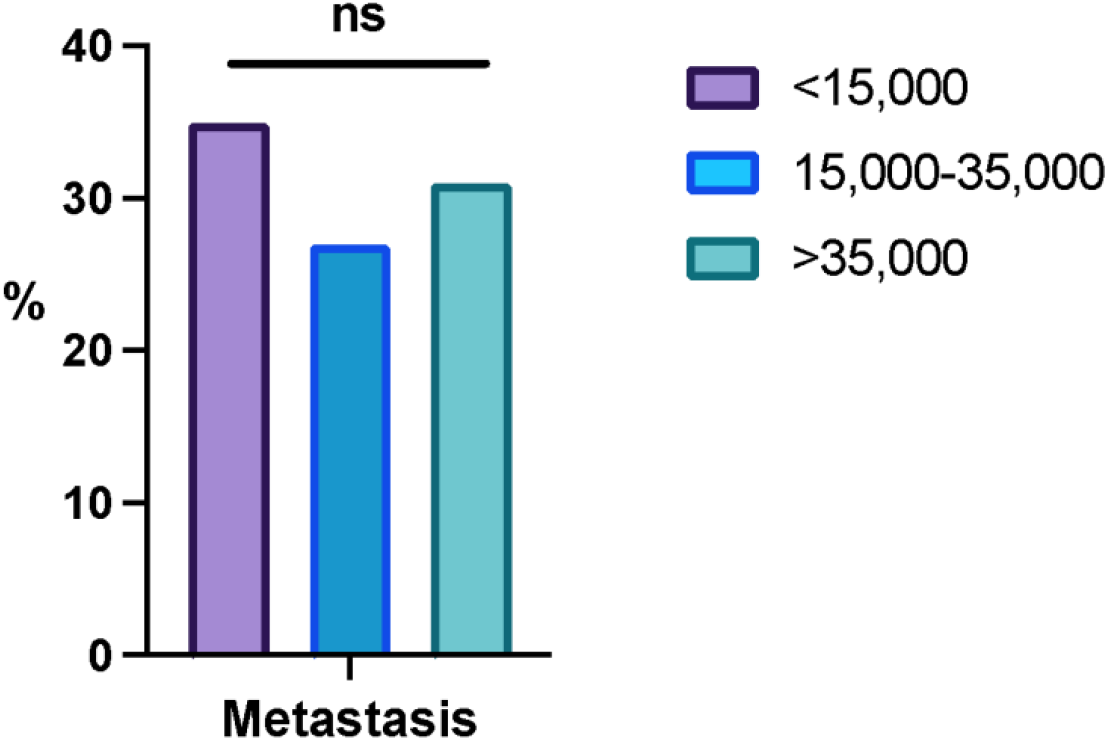
Metastatic breast cancer is similar among women in the US. We compared metastatic breast cancer rates and socioeconomic status. Three groups: less than $15,000, $15,000-$35,000, and greater than $35,000, were compared for the percentage of metastatic disease.

## Discussion

To address the underrepresentation of Latin American/Hispanic women in breast cancer genetic studies, we explored two strategies for patient recruitment: a community-based model and a hospital/cancer center-based model. For community recruitment, we designed and received approval for a protocol allowing patient ascertainment by phone, through the internet, and in person. We advertised the protocol on clinicaltrials.gov, through social media, sending letters to oncologists in predominantly Hispanic areas, personal contacts with Hispanic cancer support and business groups, and through charities. Although we reached a broad audience, only 38 patients were enrolled by this approach. We observed that in-person presentations in Spanish to breast cancer patients and survivors resulted in high participation. However, the study needed full-time staff and resources to pursue this approach on a national level. Establishing the protocol in hospitals with sizable Hispanic patient populations in the US, such as Lubbock and El Paso, Texas, as well as an adult cancer hospital and a large general hospital in Guatemala, was far more effective. Although the study did not have funds for the Texas hospitals to pay for staff time, we still enrolled 179 subjects in 7.5 years in Lubbock and 70 subjects in 0.5 years in El Paso. In Guatemala, we funded staff to recruit, consent, administer questionnaires, collect, and ship samples, and this yielded 685 subjects in 5.25 years at INCAN and 116 subjects in 3.5 years at HGSJDD. We conclude that community-based recruitment can be effective, but to be successful requires in-person contact with patients and a dedicated bilingual staff. Hospital-based studies in areas with a substantial Hispanic population are also effective, if adequate staff is available.

Large patient studies analyzing the participation of minorities in genetic studies found the participation of Hispanic women was 73-87% [4]. The recruitment strategies for these studies included telephoning patients directly and providing transportation for biospecimen collection.

Although these studies demonstrated effective community-based recruitment, the authors emphasized the need for substantial financial support for extensive genetic studies [4, 11].

### Genetic testing

Genetic testing is largely unavailable in most low- and middle-income countries like Guatemala, resulting in higher morbidity and mortality. In addition, genetic testing is underutilized in US Hispanic populations due to a lack of insurance and understanding the value of genetic testing. However, the increase of Latin American women in genetic studies improves health outcomes. Our data shows that 10% of Latin American unselected women have a germline pathogenic mutation. Furthermore, similar to other populations, Latin American mutation carriers have an earlier age of onset, triple-negative disease, and relevant family history.

Our current data on pathogenic mutations extend results we previously published on these patient populations [5, 8], finding additional examples of the 212+1 G>A founder mutation in Guatemala [12]. We also identified one case from Texas carrying the c.68_69delAG/185delAG mutation. This mutation is prevalent among women of Ashkenazi-Jewish descent [13]. In previous studies, 185delAG was seen in specific Latin American and Mexican American populations as the most common mutation [14, 15]. The 185delAG carrier we identified had triple-negative breast cancer with metastasis, was diagnosed at 40-44 years of age, and had first- and second-degree relatives with breast cancer. Women with pathogenic mutations can benefit from enhanced screening and prophylactic surgery to prevent breast and ovarian cancer.

Therefore, enhanced use of genetic testing and counseling would result in earlier diagnosis and improve outcomes across Hispanic communities in the US and Latin America.

Genome-wide association studies (GWAS) of breast cancer have identified over 170 loci [16] associated with this disease. Interestingly, a GWAS study in 1,497 Latina women in the US identified a protective variant (rs140068132) on chromosome 6q25. This variant is found almost exclusively in people of Indigenous American ancestry. Furthermore, the variant is associated with lower mammographic density, reduced ER-negative cancer, and reduced overall breast cancer risk (16%). This study further emphasizes the importance of genetic studies in Latin American women [17].

### Triple-negative breast cancer

Breast cancer subtyping is critical to determining therapy and outcomes. For example, triple-negative breast cancer is associated with a higher rate of metastasis, lower response to therapy, and lower survival rates than other breast cancer subtypes [18, 19].

TNBC prevalence varies by ethnicity, with approximately 10% in women of European descent, 30% in African American women, and 10-20% in Hispanic women [20-22]. In our study, 17% of US Hispanic women had triple-negative breast cancer. Women with germline mutations had a significantly higher frequency (41%) of TNBC, than in women without (15%) (P=0.01). *BRCA1* and to a lesser extent, *BRCA2* mutations were associated with TNBC, as seen in other populations [23].

## Limitations

The limitations of this study include the use of self-reported data in the questionnaire, and that information on treatment and outcomes was not available. In Guatemalan women, direct SES data were unavailable; therefore, we used cooking with wood to ascertain women with a low SES and gas stove cooking for higher SES. In addition, the sample size of the community-recruited US Hispanic women was modest.

## Summary

There is limited representation of Latin American women in genetic studies and extensive epidemiology studies rely on hospital-based databases. Hospital-based recruitment proved more productive due to the continuity of communication and recordkeeping, and community recruitment requires substantial financial support.

In both the US and Guatemala, women with pathogenic mutations have a significantly younger age of diagnosis. In addition, they are more likely to have a first- or second-degree relative with breast cancer. Also, women with a pathogenic variant were more likely to develop triple-negative disease. Our results emphasize the importance of the availability of genetic testing for all women with breast cancer.

## Data Availability

All data produced in the present study are available upon reasonable request to the authors.

## Acknowledgements

We thank Lineth Boror, Ester Avila, and Patricia Zaid for sample collection. The authors acknowledge the research contributions of the Cancer Genomics Research Laboratory for their expertise, execution, and support of this research in the areas of project planning, wet laboratory processing of specimens, and bioinformatics analysis of generated data. The content of this publication does not necessarily reflect the views or policies of the Department of Health and Human Services, nor does mention of trade names, commercial products, or organizations imply endorsement by the U.S. Government. We are grateful for the use of the NIH Helix Biowulf computing facility.

## Funding

This project has been funded in whole or in part with Federal funds from the National Cancer Institute, National Institutes of Health, under NCI Contract No. 75N910D00024.

## Competing Interests

The authors have no relevant financial or non-financial interests to disclose.

## Author Contributions

Michael Dean contributed to the study conception and design. Material preparation, data collection and analysis were performed by Roberto Orozco, Eduardo Gharzouzi, Michael Dean The first draft of the manuscript was written by Jesica Godinez Paredes, Isabel Rodriguez, and Michael Dean. All authors read and approved the final manuscript.

## Data Availability

The datasets generated during and/or analyzed during the current study are available in the dbGAP repository, dbGaP Study Accession: phs002246.v1.p1.

## Ethics approval

The study was approved by the Institutional review Board of the NCI. ClinicalTrials.gov Identifier: NCT01251900) and the research committees of the Instituto de Cancerologia and the Guatemalan Ministry of Health.

## Consent to participate

Informed consent was obtained from all individual participants included in the study.

## Notes

### Competing Interest Statement

The authors have declared no competing interest.

### Clinical Protocols

https://classic.clinicaltrials.gov/ct2/show/NCT01251900

### Author Declarations

The Institutional Review Board of the National Cancer Institute approved the study. ClinicalTrials.gov Identifier: NCT01251900) and the research committees of the Instituto de Cancerologia and the Guatemalan Ministry of Health.

